# Refining the Diagnostic Accuracy of Parkinsonian Disorders using Metaphenomic Annotation of the Clinicopathological Literature

**DOI:** 10.1101/2023.12.12.23299891

**Authors:** Quin Massey, Leonidas Nihoyannopoulos, Peter Zeidman, Tom Warner, Kailash Bhatia, Sonia Gandhi, Christian Lambert

## Abstract

**Background:** The diagnostic precision of Parkinsonian disorders is not accurate enough. Even in expert clinics up to one in five diagnoses are incorrect. This leads to cohorts with mixed pathologies, impacting our ability to understand disease heterogeneity and posing a major challenge for clinical trials. Gold standard diagnosis is post-mortem confirmation of the underlying proteinopathy, however many clinicopathological studies focus on either a single disease or frame analyses in one temporal direction (i.e., in-life diagnosis vs post-mortem or vice versa). Given Parkinson’s Disease (PD), Multiple System Atrophy (MSA), Progressive Supranuclear Gaze Palsy (PSP), Dementia with Lewy Bodies (DLB) and Corticobasal degeneration (CBD) can all mimic one-another, these may underestimate mis- and missed diagnoses.

**Methods:** The objective was to comprehensively map the mis- and missed diagnoses across the Parkinsonian disorders and use phenotypic features to develop a probabilistic model to refine diagnostic likelihoods based on clinical observations. We identified 125 published clinicopathological cohorts and case-reports since 1992, extracted phenotype information for ∼9200 post-mortem cases, and curated the data in a standardized machine-readable format.

**Findings:** MSA diagnostic accuracy was highest (92·8%) and DLB lowest (82·1%). MSA and PSP were most frequently mis-labelled as PD in life (7·2% and 8·3% of cases), where-as the most common PD misdiagnosis was Alzheimer’s (∼7% cases). DLB age at diagnosis was older, CBD younger, and survival longer in PD. Clinical annotation was extremely variable, which represents a limitation with clinicopathological literature, however we created likelihood ratios for a range of features and demonstrate how these can refine diagnoses.

**Interpretation:** This work delivers a harmonized, open-source dataset representing over 30 years of published results and represents a key foundation for more flexible predictive models that leverage different sources of information to better discriminate Parkinsonian disorders during the early and prodromal phases of the illness.

**Funding:** Medical Research Council

**Research in context:** *Evidence before this study:* The diagnostic precision of Parkinsonian disorders is not accurate enough – estimated misdiagnosis rates, derived from clinicopathological studies, vary between 10 – 20% depending on the condition, context and criteria. However, many previous studies either focus on one single condition, or frame the analysis in one temporal direction. By the time Parkinsonian disorders manifest with motor symptoms, the conditions have been present for 10-20y. Previous work has proposed a probabilistic approach to identify prodromal Parkinson’s disease, but none exist for the range of common Parkinsonian disorders that often mimic one another.

*Added value of this study:* This study structures and standardises 30-years of clinicopathological data across all the main Parkinsonian syndromes, making it available in an open, machine-readable format, and also updates the Human Phenotyping Ontology for Parkinsonian syndromes. It uses these to comprehensively map the patterns of missed and mis-diagnosis across all of the conditions, and build a flexible multimodal probabilistic approach to help refine diagnoses of these disorders.

*Implications of all the available evidence:* This work provides a key foundation for a modular framework that can be flexibly adapted and combined with different tools, techniques and approaches to more accurately diagnose different Parkinsonian disorders during the early and prodromal phases of the illness.

## 1. INTRODUCTION

Parkinson’s disease (PD) is the second most common neurodegenerative illness. It presents as a motor syndrome that emerges once 60-70% of the nigral dopaminergic neurons have been irreversibly lost. ^1^ Diagnosis is clinical based on the cardinal signs of bradykinesia with rigidity and/or tremor, coupled with a lack of features to indicate an atypical Parkinsonian syndrome (aPD). ^1^ Once diagnosed, progression is highly variable, with survival ranging from a few years to several decades. ^2^

The diagnostic precision of Parkinsonian disorders is not accurate enough. Even in expert clinics up to one in five PD diagnoses are incorrect. ^3^ aPD conditions are common mimics, which include Multiple System Atrophy (MSA), Progressive Supranuclear Palsy (PSP), dementia with Lewy bodies (DLB) and Corticobasal Degeneration (CBD). ^3^ Furthermore, approximately 50% of the more aggressive forms of PD, the so-called *malignant* phenotype, are mis-diagnosed as aPD in life. ^4^ These represent a challenge for developing disease modifying treatments, as clinical trial cohorts will contain mixtures of pathologies (*misdiagnoses*) necessitating larger sample sizes to detect a signal, and subtypes of disease with markedly different disease trajectories, that may require more aggressive or targeted therapies, ^5^ will be underrepresented (*missed diagnoses*).

The diagnostic gold standard for these disorders is post-mortem confirmation of the underlying proteinopathy. However, many clinicopathological studies focus on either a single diagnostic entity or frame the analyses in one temporal direction (i.e., life diagnosis vs post-mortem findings or visa versa). Given that each of the Parkinsonian disorders can mimic one-another, these risk missing the true extent of mis- and missed diagnoses. The value of pathologically confirmed cases is high, and there is a wealth of data embedded in historic reports that could be leveraged to help improve diagnostic precision in life.

Phenotypes are defined as *“any observable characteristic of an organism”* ^6^ and therefore span many body systems and multiple levels of scale. Often, the terminologies used to define different observations vary between experimenters, making systematic comparisons hard. To tackle this complexity, ontologies seek to formalise and structure the language used to describe different observations, making them more suitable for large scale computational analyses and comparisons. ^7^ In human disease, the Human Phenotyping Ontology (HPO) is a highly successful framework for deep phenotyping (https://hpo.jax.org/app/). Whilst there has been an independent Parkinson’s disease ontology (PDON), ^8^ HPO is actively maintained, regularly updated through community feedback to iterate and refine, and has been adopted by large initiatives such as the 100,000 Genomes Project.

*Metaphenomic annotation* is a novel method to structure data from published cohorts or single case reports in a standardised, machine-readable format based around internationally recognised phenotyping ontologies and structures, leveraging the Phenopacket standard for structuring phenotype data (http://phenopackets.org). It allows more efficient pooling of phenotyping data that can then be used for a wide array of different analyses.

In this work, we used metaphenomic annotation on the clinicopathological literature for Parkinsonian disorders published since the 1992 validation of the Queen Square Brain Bank Criteria for PD. ^9^ The objective was to comprehensively map the mis- and missed diagnoses across the main Parkinsonian disorders and link these gold-standard cases to the phenotypic features observed in life. These results form the foundation for a naïve Bayesian classifier, ^1^ that can be used to quantify the probability of disease for each of the main Parkinsonian syndromes. These results can be flexibly expanded or incorporated into other tools, modalities or risk scores seeking to improve the diagnostic accuracy across the Parkinsonian disorders, and deliver a freely accessible, machine-readable library summarising the last 30-years of published data.

## 2. METHODS

### 2.1 Literature Review

A Pubmed search was performed between the dates 1/9/1992 – 1/12/2022 using the keywords *“Post-mortem”* or *“Clinical-pathological”* combined with: *“Parkinson’s disease”, “Dementia with Lewy Bodies”, “Multiple system atrophy”, “Progressive Supranuclear Palsy”, “Cortico-basal degeneration”, Parkinsonism”*. 663 unique articles were identified and reviewed (QM, LN, CL). Exclusion criteria included: 1. No post-mortem data; 2. Monogenic disease; 3. No data for main Parkinsonian conditions; 4. Review articles; 5. Not available in English. 6. No basic diagnostic data (i.e., age at diagnosis or disease duration); 7. Unable to annotate (e.g., no extractable data or complex mixed phenotypes). In total, 125 publications (Supplementary data) were annotated and used for analysis (Figure 1).

**Figure 1:**
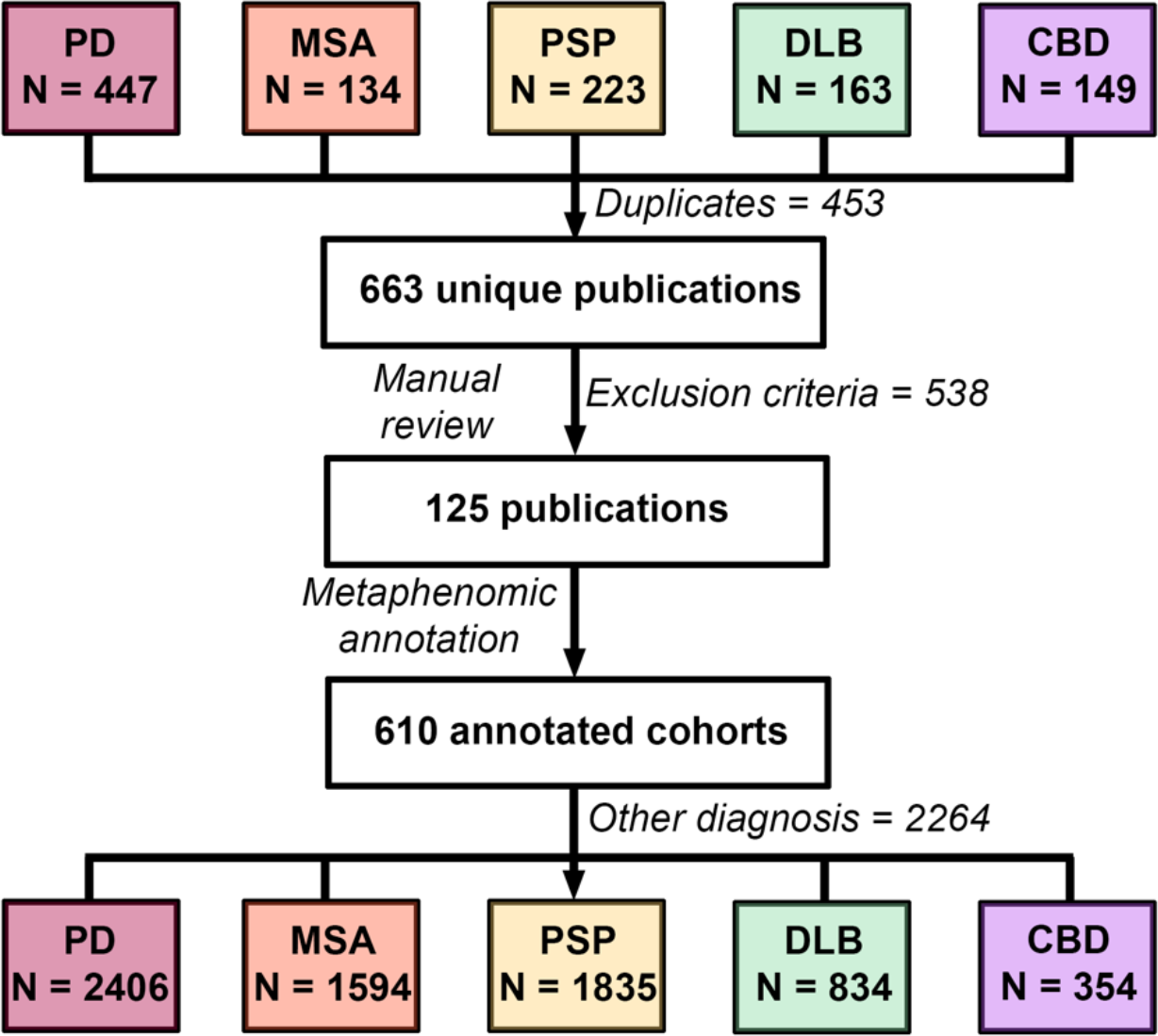
Data generation: Pubmed search between the dates 1/9/1992 – 1/12/2022 using the keywords *“Post-mortem”* or *“Clinical-pathological”* combined with each condition shown. Top row shows number of publications and filtering process. Bottom row show the total number of cases in each cohort generated through the metaphenomic annotation. Note, of these 9287 cases only 5748 provided misdiagnosis data as detailed in the main text.

### 2.2 Metaphenomic annotation

Phenopackets (http://phenopackets.org) is a proposed standard for structuring and sharing disease and phenotype data. However, it has primarily been designed for in-person assessment of single cases. Metaphenomic annotation, introduced here, adapts this framework for published phenotyping data, both single subjects and cohorts, following the recommended best practice (see supplementary data). It is implemented as a freely available MATLAB toolbox (url) for the Statistical Parametric Mapping software (SPM, https://www.fil.ion.ucl.ac.uk/spm/software/spm12/).

### 2.3 Phenotype and Disease Ontologies

The “*Human Phenotyping Ontology*” (HPO) and MONDO library of human disease was used throughout. To ensure adequate coverage we transcribed all clinical phenotype terms from the PD Ontology ^8^ to HPO. Any absent terms identified through this work were defined and submitted to the HPO team to provide better coverage.

### 2.4 Analysis

For all individuals with a diagnosis of sporadic PD, MSA, PSP, DLB and CBD we extracted: age of onset (symptom where available, or diagnosis if unavailable), age at death, phenotypes, misdiagnosis data, disease duration. At post-mortem, it is not possible to separate DLB from PD Dementia, and several studies subsume these as “Lewy Body Disease”. Here, we only present results from studies defining PD and DLB as separate cohorts, however “Lewy Body Disease” results are available via our analysis code. Misdiagnoses falling outside these main disorders were classified as “OTHER”. The misdiagnosis analyses excluded cohort studies that did not report this feature.

Analysis was done in MATLAB 2021b. Summary statistics were combined using pooled variance and mean. ^10^ For each dataset, the ratio between sample size versus the total final number in each cohort was used to calculate weighted means. Cohort differences in onset, disease duration and age of death were tested using a Krushkal-Wallis test. If significant, Wilcoxon rank sum test was used for pair-wise comparisons (Bonferroni P < 0·005). To summarise misdiagnosis data, we collapsed each disease into a 2 × 2 confusion matrix where diagnosis in life was framed as the prediction and pathological diagnosis ground truth. In this way sensitivity, specificity and balanced accuracy were calculated for each diagnosis conditional on the other disorders (detailed in supplementary data).

### 2.5 Probability of disease from phenotypic features

We used a naïve Bayesian classifier approach similar to that proposed for prodromal PD. ^1^ Details on how it was adapted for this work is in the supplementary data. In the results, we provide a worked example how this can be used to refine diagnoses. For this, likelihood ratios (LRs) were calculated from the sensitivity and specificity as follows: ^12^

- Positive Likelihood Ratio = Sensitivity/(1-Specificity)
- Negative Likelihood Ratio = (1-Sensitivity)/Specificity

For the example of neurofilament light chain, the published sensitivity was 0·86 and specificity 0·85. ^11^ These were only available relative to aPD cohorts (MSA/PSP), hence we inverted them (i.e., 1 – value) to calculate values for non-aPD groups.

### 2.6 Data availability

All the annotated .json files are freely available at: [url tbc] All results of the presented analysis can be reproduced via: [url tbc].

## 3. RESULTS

### 3.1 Cohort

125 publications were identified, generating 610 annotations totaling 9287 post-mortem diagnosed cases (2406 PD, 1594 MSA, 1835 PSP, 834 DLB, 354 CBD, 2264 other), which were used for age of onset and survival analyses. Of these, 5748 provided misdiagnosis data (1698 PD, 965 MSA, 1349 PSP, 347 DLB, 265 CBD, 1124 other). The “*other*” diagnostic category was most frequently Alzheimer’s disease (86%) followed by Frontotemporal dementia (8·8%), with the PSP and CBD cases contributing significantly to the latter. This is summarized in figure 2.

**Figure 2:**
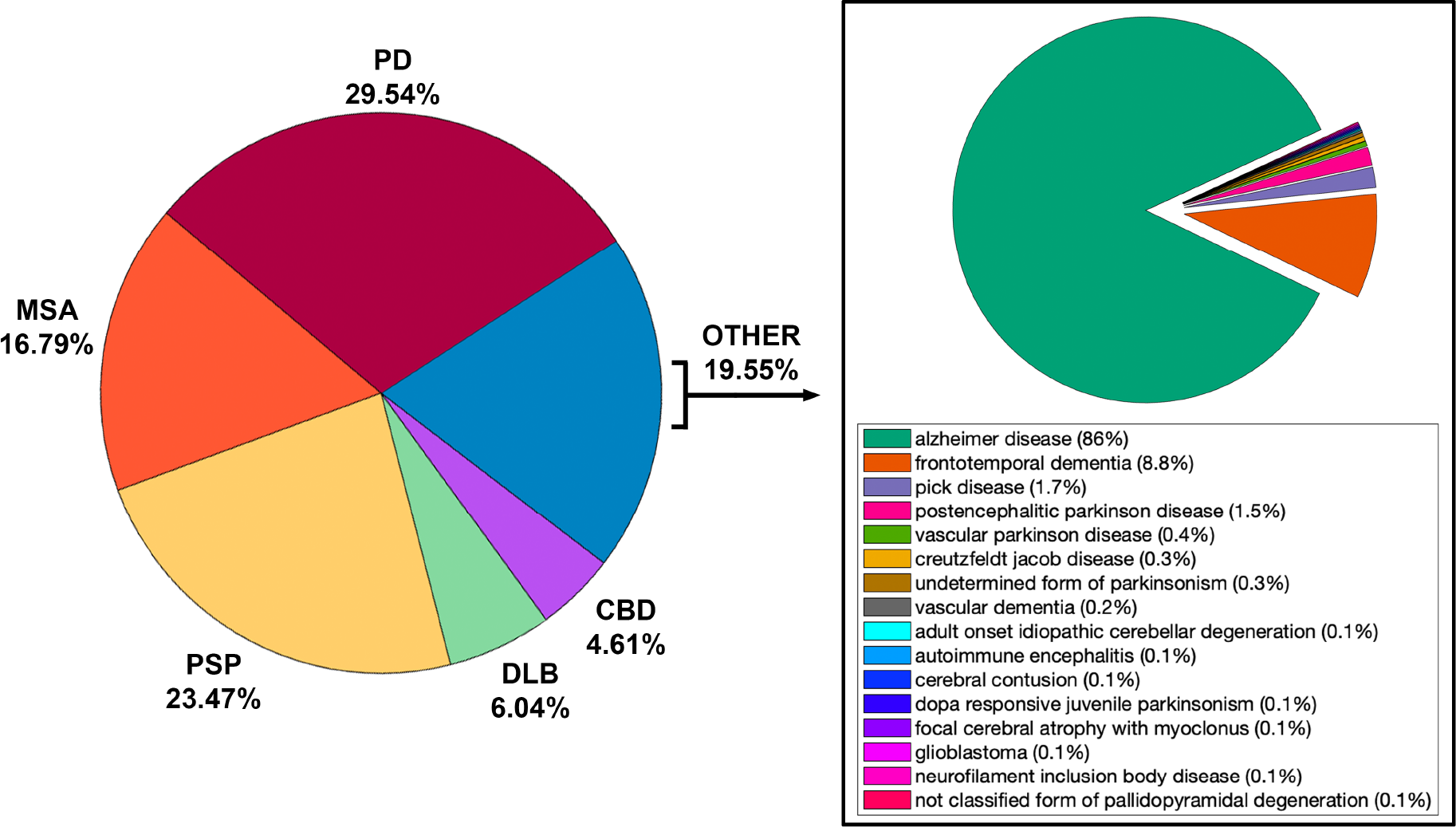
Summary of post-mortem diagnoses.

### 3.2 Age of Onset

Figure 3 summarises these results. Symptom onset for DLB was the oldest (69·34 ± 10·46y), followed by PSP (65·60 ± 8·10y), PD (62·75 ± 11·11y), CBD (62·64 ± 7·78y) and MSA the youngest (59·19 ± 9·12). DLB was significantly older than the other groups except for PD, and CBD was significantly younger except for MSA (Figure 5).

**Figure 3:**
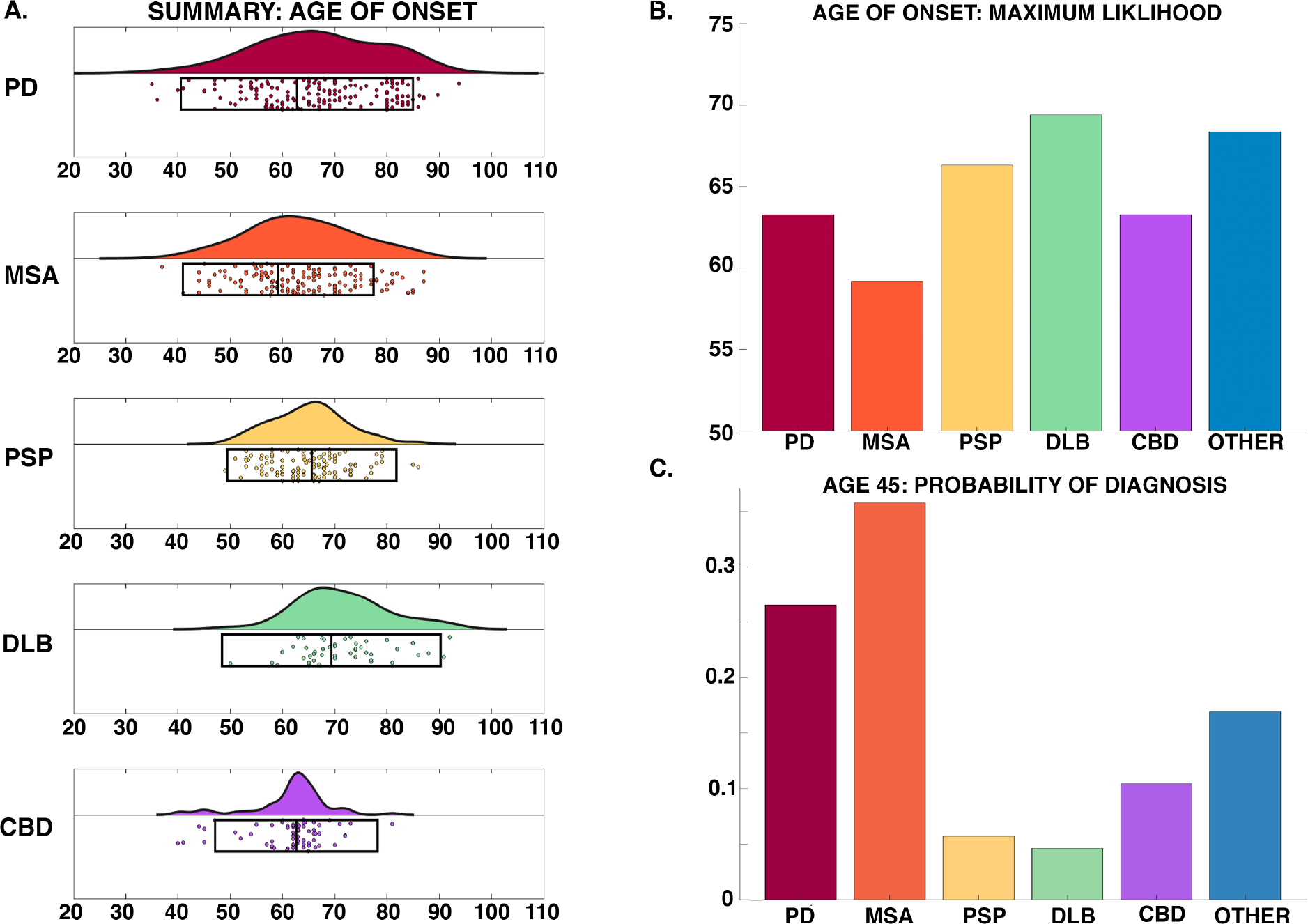
Age of onset: A. Raincloud plots summarising 9287 Parkinsonian cases – boxes below indicate weighted mean and two standard deviations. Note each scatter point may either represent a cohort study or single case reports, however these were weighted by sample size to calculate summary statistics and probabilities. No significant difference in age of onset was seen between groups; B. Age of onset maximum likelihood for each condition; C. Probability of each Parkinsonian syndrome at age 45y.

### 3.3 Survival

Figure 4 summarises the mortality data: DLB has the oldest mean age of death (78·59 ± 8·52y) followed by PD (77·37 ± 7·86y), PSP (73·87 ± 7·93), CBD (70·77± 7·64) then MSA (66·49 ± 8·52). PD and DLB were significantly older than the rest of the groups, and CBD and MSA significantly younger than PSP (Figure 5). Duration of survival for aPD was similar with MSA (7·19 ± 2·60), PSP (7·39 ± 3·80), DLB (7·85 ± 5·75) and CBD (6·91± 3·26). PD survived significantly longer (14·64 ± 6·96) with a disease duration ranging from 2 - 34 years.

**Figure 4:**
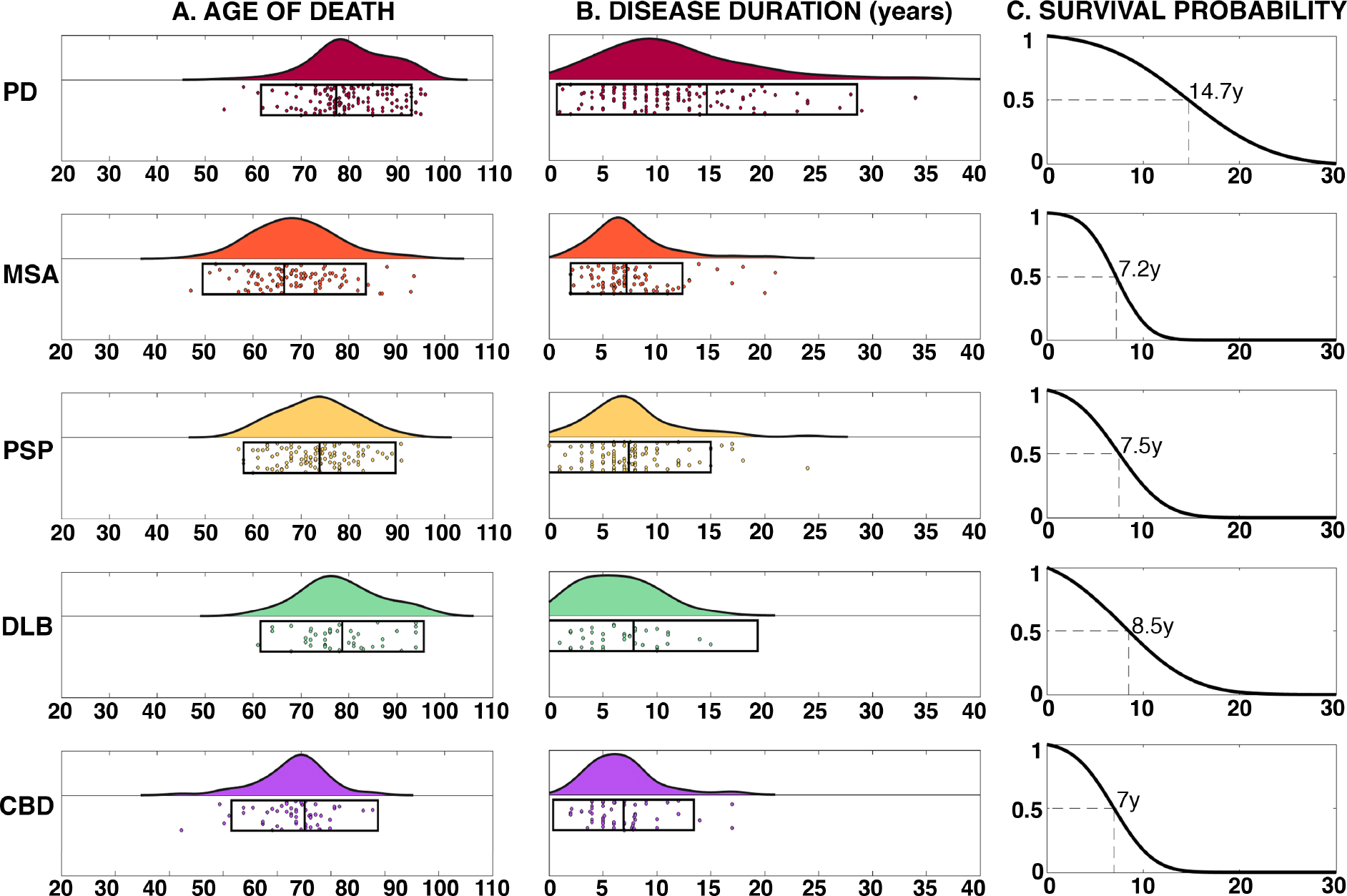
Survival: A. Raincloud plots summarising age of death for each condition; B. Raincloud plot summarizing disease duration in years; C. Cumulative probability of survival from symptom onset in years with 50% survival point labelled.

**Figure 5:**
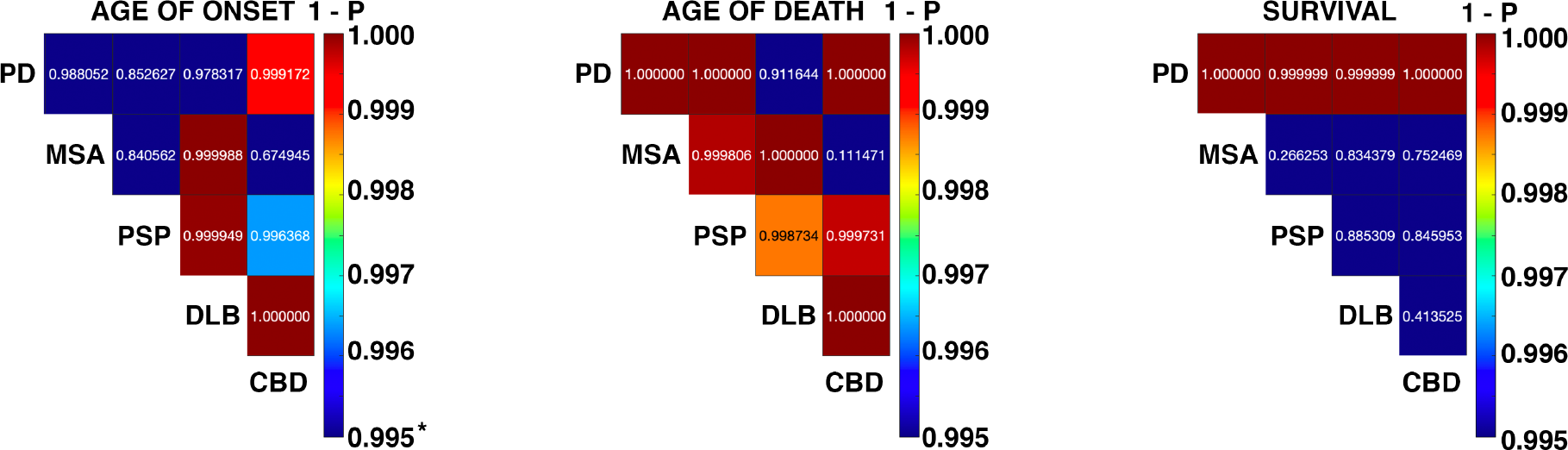
Summary of statistical tests: Heatmaps of one minus p value summarising each of the pairwise tests for age of onset, age of death and survival (disease duration), thresholded at Bonferroni corrected P < 0·005

### 3.4 Misdiagnosis

This is summarised in Figure 6 and table 1. Balanced accuracy was lowest for DLB, with a significant number labelled as MSA or OTHER (mainly AD). CBD was the next lowest, however in life this presents as corticobasal syndrome that is often due to PSP or FTD, as reflected in our results (17·38% PSP, 13·12% other). Of note, ∼5% of CBD cases are labelled as PD in life. PSP was next due to the lower sensitivity compared to PD and MSA. This was caused by in-life PSP mimics caused predominately by PD (3·01%), CBD (2·51%) and OTHER (3·68%), and cases of PSP being mislabeled as PD (8·52%), MSA (5·63%) and CBD (3·63%). The balanced accuracy for PD was ∼90%, but with comparatively large proportions of AD mimics (∼7%) in life and with ∼8% of cases misdiagnosed as aPD, most often MSA (5·42%). This latter result is in-line with observations that up to half of the more aggressive, malignant form of PD are diagnosed as aPD. ^4^ MSA was the most accurate overall with a balanced accuracy of 92·82%, and similar numbers of PD and PSP mimics in life (8·29% and 6·85% respectably). Between the more common aPD conditions, PSP and MSA, there were similar numbers being mislabeled as PD in life (7·36%, 8·52%). There is evidence in PSP these Parkinsonian variants follow a less aggressive course with longer survival, but not in MSA. ^13^

**Table 1:** Accuracy metrics pooling mis- and missed diagnoses data.

| Diagnosis | Sensitivity (%) | Specificity (%) | Balanced Accuracy (%) |
| --- | --- | --- | --- |
| PD | 87.51 | 91.95 | 89.73 |
| MSA | 90.57 | 95.07 | 92.82 |
| PSP | 79.32 | 97.16 | 88.24 |
| DLB | 64.84 | 99.26 | 82.05 |
| CBD | 71.70 | 98.32 | 85.01 |
| OTHER | 75.89 | 95.00 | 85.45 |

**Figure 6:**
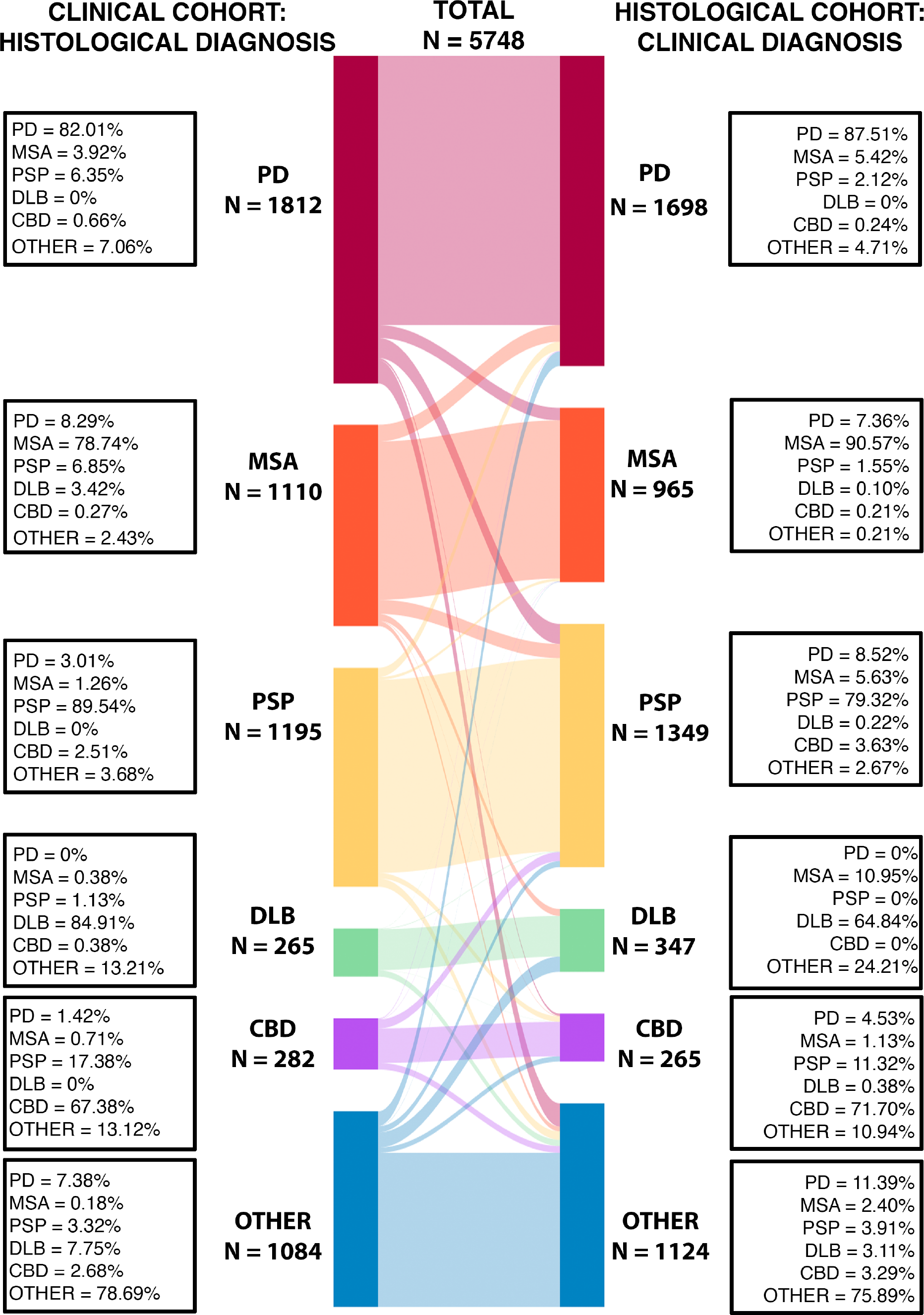
Misdiagnosis (left, clinical diagnosis mapped to post-mortem) and missed diagnosis (right, post-mortem mapped to clinical label) between conditions.

### 3.5 Improving Diagnostic Accuracy using Phenotypic Data

88% of studies had extractable phenotyping data providing 4076 descriptors. These were mapped to 246 unique HPO descriptors over 12 parent domains. From this HPO graph, we calculated the top five phenotype terms with the largest likelihood ratios between diseases, reflecting clinical observations that can co-occur in both diseases and best discriminate between the two (figure 7). This unbiased approach confirms certain highly predictive clinical features such as pill-rolling/rest tremors in PD, ataxia and stridor in MSA, cognitive impairment DLB, gaze palsies and falls PSP, and cortical sensory loss CBD (Figure 8). This analysis excluded empty observations, which may pathognomonic signs for one disease versus another, because we could not assume absent reporting equated to an absent sign. Relaxing this constraint confirms this assumption. For example, cortical sensory loss has never been reported in PD, MSA and DLB, alien hand phenomena in MSA or DLB, and pill-rolling tremor in PSP, but then inability to walk has not been recorded in DLB which is clearly an artifact.

**Figure 7:**
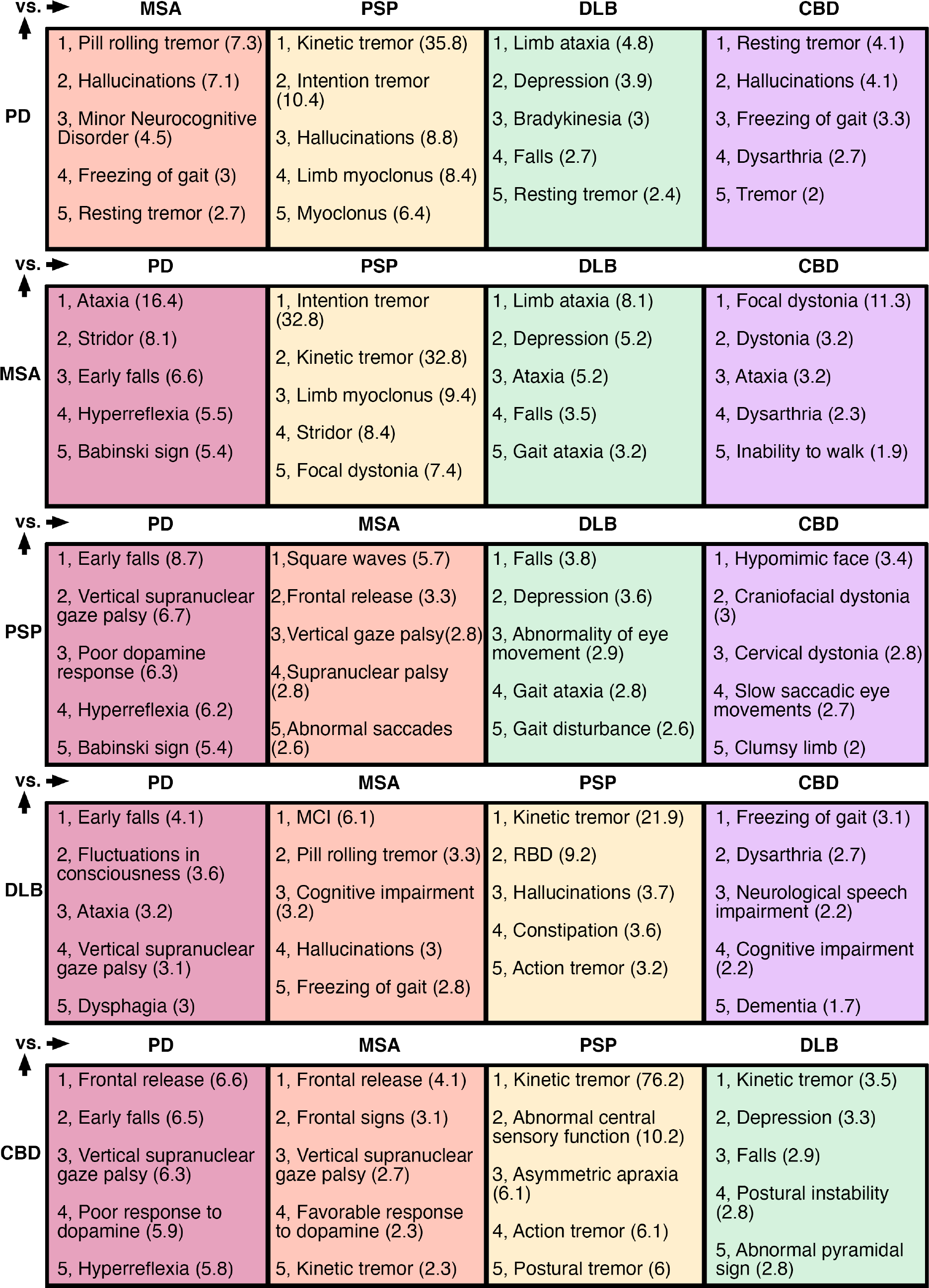
Over-lapping phenotypes with the maximum likelihood ratio over the entire HPO tree for discriminating the condition on the left, from each of the main mimics (top row). Positive likelihood ratio provided in the brackets. Abbreviations: MCI = Mild cognitive impairment. RBD = REM sleep behaviour disorder. Most of the terms are as per HPO definitions, but a few were abridged due to space constraints

Despite the limitations of incomplete reporting in post-mortem literature, providing a robust link between phenotype and pathological diagnosis provides a foundation that can be developed to improve diagnostic accuracy *in vivo*, as shown in the following example:

A 50-year-old person presents with Parkinsonism, REM Behaviour Sleep Disorder (RBD), orthostatic hypotension ^4^ and tremor. Leveraging the metaphenomic structured data we can calculate the most likely diagnosis is PD (probability = 0·97) or DLB (0·73) followed by MSA (0·58). The strength of this approach is it can easily incorporate new data to refine the prediction. If subsequent testing revealed an elevated neurofilament light chain level, ^11^ updating the calculation with the corresponding likelihood ratios would result in probabilities of 0·89 for MSA and 0·83 for PD (table 2). If, on re-examination, a rest tremor was found then PD would remain the most likely even with elevated NFLC (PD 0·99 vs MSA 0·78), but an additional history of erectile dysfunction would then make MSA more likely (MSA 0·99 vs PD 0·88).

**Table 2:**
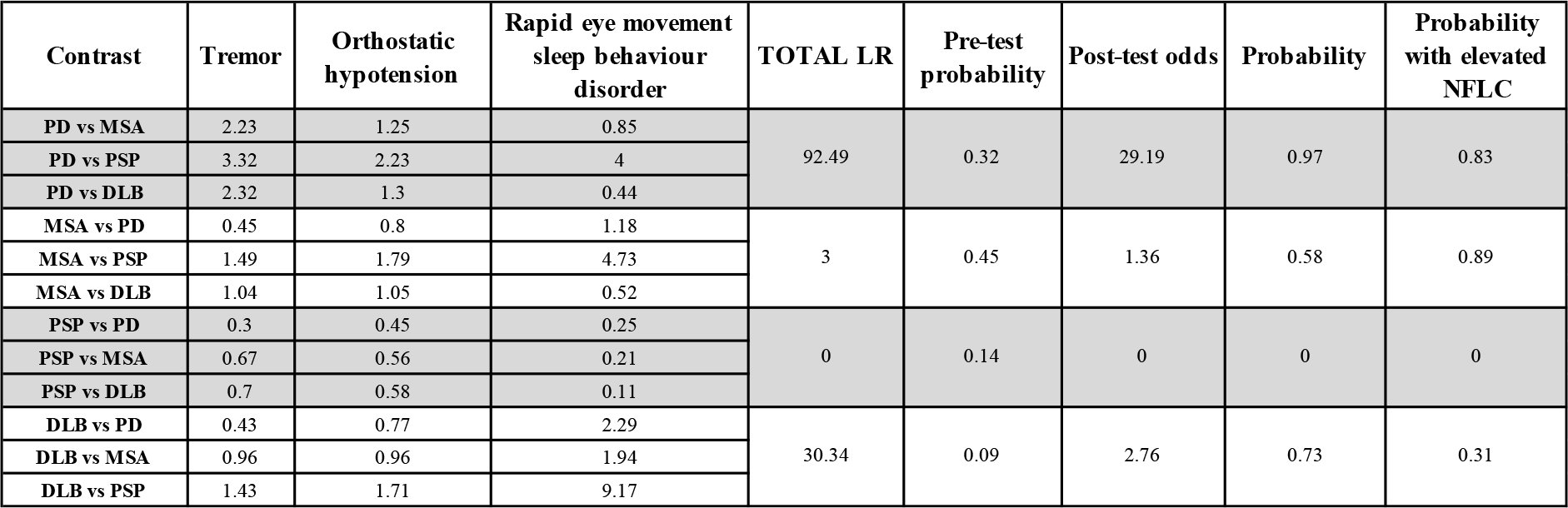
Worked example of a 50yo with Parkinsonism plus RBD, Orthostatic Hypotension and tremor. The most likely diagnosis with this combination is PD (0·97) or DLB (0·73) followed by MSA (0·58). However, if this individual has an elevated NFLC result this flips making a diagnosis of MSA more likely (0·89 vs 0·83). Ascertaining additional clinical signs enables one to refine this further – Building on the example above, if a rest tremor was present, PD would remain the most likely even with elevated NFLC (PD 0·99 vs MSA 0·78), however the additional presence of erectile dysfunction with the rest tremor then makes MSA far more likely (MSA 0·99 vs PD 0·88). Note CBD not shown as RBD has not been reported in this condition.

| Contrast | Tremor | Orthostatic hypotension | Rapid eye movement sleep behaviour disorder | TOTAL LR | Pre-test probability | Post-test odds | Probability | Probability with elevated NFLC |
| --- | --- | --- | --- | --- | --- | --- | --- | --- |
| PD vs MSA | 2.23 | 1.25 | 0.85 | 92.49 | 0.32 | 29.19 | 0.97 | 0.83 |
| PD vs PSP | 3.32 | 2.23 | 4 |  |  |  |  |  |
| PD vs DLB | 2.32 | 1.3 | 0.44 |  |  |  |  |  |
| MSA vs PD | 0.45 | 0.8 | 1.18 | 3 | 0.45 | 1.36 | 0.58 | 0.89 |
| MSA vs PSP | 1.49 | 1.79 | 4.73 |  |  |  |  |  |
| MSA vs DLB | 1.04 | 1.05 | 0.52 |  |  |  |  |  |
| PSP vs PD | 0.3 | 0.45 | 0.25 | 0 | 0.14 | 0 | 0 | 0 |
| PSP vs MSA | 0.67 | 0.56 | 0.21 |  |  |  |  |  |
| PSP vs DLB | 0.7 | 0.58 | 0.11 |  |  |  |  |  |
| DLB vs PD | 0.43 | 0.77 | 2.29 | 30.34 | 0.09 | 2.76 | 0.73 | 0.31 |
| DLB vs MSA | 0.96 | 0.96 | 1.94 |  |  |  |  |  |
| DLB vs PSP | 1.43 | 1.71 | 9.17 |  |  |  |  |  |

## 4.0 Discussion

This work structures 30 years of clinic-pathological literature for the main Parkinsonian syndromes into an easy to use, machine readable format for complex phenomic data. It establishes a foundation for clinical observations within a probabilistic diagnostic framework that underwrites transportability by design by allowing a “*plug and play*” approach where different combinations of techniques can be used and adapted to local resources. Importantly, such an approach provides quantitative metrics that are directly comparable irrespective of the combination of methods used and implicitly accounts for the underlying uncertainty inherent in all *in vivo* diagnostics.

### 4.1 Demographics

Onset ages were in keeping with existing literature, ^14^ with MSA more likely at a younger age versus PSP and DLB that tended to be later. The range was substantial, particularly in PD and MSA where it spans nearly six decades (35y - 94y). Survival in aPD was approximately 7 years, with no differences between groups. This falls within the expected range for PSP and MSA, ^15^ but for DLB was at the upper end of the expected 1·9 – 6·3y. ^16^ This discrepancy may emerge for several reasons: DLB can be a challenging to disambiguate from AD in life, ^17^ as hallucinations and Parkinsonism may occur in both, ^18^ and at post-mortem a significant proportion have dual pathology associated with more rapid progression and shorter survival. ^19^ Furthermore, DLB and PD dementia look identical at post-mortem, and the distinction hinges upon the sequence and timing of clinical events. As such, these cases are relatively under-represented in the literature (Figure 2), and the overlap with AD may bias current cohorts. PD was associated with a longer survival (14·64 ± 6·96y) consistent with previous reports, ^20^ ranging from 1 to 34 years. The reasons for this heterogeneity are unknown, with age of onset, akinetic phenotype, cognitive dysfunction and GBA1 gene variants all associated with more rapid disease progression, ^20^ where-as lifestyle factors such as physical exercise seem to exert protective effects.

### 4.2 Misdiagnoses and Missed diagnoses in Parkinsonian Syndromes

The diagnostic accuracy of Parkinsonian disorders remains suboptimal, and partly dependent on clinician experience, years from symptom onset and clinical phenotype. ^14^ For PD, balanced accuracy was 89·7%, in line with Adler and colleagues. ^21^ For MSA it was >90%, higher than the expected 70-80%. ^22^ Reasons for this may include taking the final diagnosis at death, greater sample size and improvements in the diagnostic criteria over time. ^23^ Accuracy for DLB was one of the lowest, with many cases mis-labelled either as AD or MSA. As noted, AD co-pathology is common in DLB which may make the distinction tricky. The presence of autonomic failure is likely to account for the confusion with MSA. Whilst cognitive impairment was considered atypical for MSA it has become increasingly recognized, ^24^ although it occurs later in the disease. In line with previous work, AD was the most frequent “*other*” diagnosis across all conditions, particularly LBD. Parkinsonism is common in AD, ^25^ with similar dopaminergic cell loss associated with regional neurofibrillary tangles. ^25^

### 4.3 Improving Diagnostic Accuracy in Parkinsonian Syndromes

The prodromal phase of Parkinsonian disorders ranges between 5 – 20 years.^1,26^ Identifying individuals during this period is critical for disease modifying therapies. However, a key factor is how accurately can we identify these conditions and discriminate them from potential mimics? Given the substantial heterogeneity within clinic-pathological defined cohorts, this remains a major challenge. It is unlikely that there will be one “*best test*” that will work across all disorders, is universally available and feasible in all scenarios. More likely, a tactical combination of investigations combined with clinical knowledge will need to be applied at the individual subject level, ^27^ and there will be a trade-off between how much new information each test provides, how invasive the procedure is, patient choice, availability and cost. For example, idiopathic anosmia is a risk factor for PD, with 1 in 10 individuals later developing the condition. There is a new CSF RTquic test to detect abnormal alpha synuclein aggregation with a sensitivity of 98% and specificity of 95·3. ^28^ Assume we want to use this to diagnose pre-motor LBD. If 10,000 anosmics are tested, 1,000 will have early LBD, and this test will detect 980 of them (sensitivity 98%). However, 9,000 will not have LBD but 432 individuals will have a positive test (i.e., specificity of 95·3) meaning ∼50% of the positive diagnostic tests will not have LBD.

The probabilistic approach to diagnosis and stratification offers a powerful framework to flexibly combine different techniques and boost diagnostic accuracy, without having to commit to one single approach, test or method, thereby providing something more universally applicable across different healthcare systems and scenarios. If the same example of anosmia is viewed in terms of probabilities (here is 0·01, and LR+ for RTquic is 20·41), a positive test alone equate to a 0·16 probability of prodromal LBD. Viewed in this light, a higher degree of confidence would be warranted before committing to a diagnosis and life-long treatment. This can be achieved through some simple additional details. ^27,29^ If we add the stipulation that they are all older than 60, the probability increases to 0·52, with additional features such as subtle motor abnormalities and an affected relative boosting it to 0·91 (vs <0·001 if anosmic over 60 where these other features are absent). ^29^ This process can also be used in reverse, to identify the most informative tests, investigations or clinical findings that would best help reconcile a diagnostic dilemma and provide an upper bound on the degree of confidence that can be achieved via different approaches. Finally, it could be used to refine diagnostic criteria, which currently rely on a step-wise, categorical approach to try and achieve the right balance between sensitivity and specificity, but invariable results in missed cases. The diagnostic guidelines for CBD provide a good example: ^30^ Rest tremor is currently an absolute exclusion, but in this work was present in 14% of post-mortem cases that assessed tremor (N = 132) and ∼5% CBD cases were mislabeled as PD in life. Viewing this same information as probabilities reveals the likelihood of CBD with rest tremor at the age of 50 is ∼2%, compared to ∼96% PD, that could then be modified by the presence/absence of other clinical features or biomarkers.

### 4.4 Limitations

There are several limitations with this current work. Whilst every effort was made to review and annotate all available literature, some could not be obtained or were not in English. However, given the overall numbers we do not feel this will significantly impact the result. Furthermore, certain cohorts were difficult to classify using the initial framework, specifically those with dual diagnoses. Whilst in the annotation we included a separate *“dual diagnosis”* category if this was clearly identifiable and extracted histological staging data, the distinction between two diseases versus low-level mixed proteinopathies is not well defined nor historically reported, and represents an open challenge. There was marked heterogeneity in precisely what was reported in the literature in terms of diagnostic milestones, demographics and phenotypic features. Regarding the latter, it had to be assumed that a lack of reporting did not equate to absent signs, which had the problem of conflating publication bias for rarer features. We attempted to factor for this by imposing a minimum number of observations, but moving forward another option would be to repeat this approach with clinical data from large cohort studies in life, where we can quantify the probability of misdiagnosis using the data collected here to better combine the two. This deeper coverage may also allow the inclusion of “*pathognomonic”* phenotypes (i.e., those with an infinite LR), which we excluded from this work due to the patchy and inconsistent reporting in published reports. Finally, whilst this work highlights the power of leveraging existing, large cohorts and data to help develop new tools to refine and improve diagnostic accuracies, it also reveals the stark limitations caused by the lack of standardisation across disciplines, specialists and journals for reporting and describing neurological cohorts. Establishing a commonly agreed framework, such as the phenopacket framework, would rapidly deliver significant gains and provide resources to better understand these complex diseases.

## 5.0 Conclusion

We have used metaphenomic annotation to structure and standardise 30-years of clinic-pathological data in Parkinsonian syndromes. We have made these resources freely available (url tbc) in addition to the full codebase to reproduce the entire analysis presented here (url tbc). We have used this to begin to build a probabilistic approach to quantify and refine diagnostic precision across Parkinsonian syndromes, providing a foundation for a modular framework that can be flexibly adapted and combined with different tools, techniques and approaches to more accurately diagnose different Parkinsonian disorders during the early and prodromal phases of the illness.

## Contributors

CL designed the study, created the annotation software and designed the methodology. QM and CL did the literature search and collected the data with help LN. CL and QM did the analysis, code and figures. CL, QM and LN collated missing HPO terms, and defined these with input from SG, KB, and submitted them to HPO. All authors contributed to the interpretation of the results. QM and CL wrote the original draft of the manuscript and SG, KB, TW, PZ and LN reviewed and critically revised the manuscript. All authors approved the final version for submission and accept responsibility for submitting for publication.

## Acknowledgements

CL was supported by an MRC Clinician Scientist award (MR/R006504/1). The Wellcome Centre for Human Neuroimaging is supported by core funding from the Wellcome Trust (203147/Z/16/Z)

## Supplementary Material

**S1.1 Adapting Phenopackets for Metaphenomic Annotation**

The basic Phenopackets ^1^ structure uses a protobuf schema which is “*a language-neutral, platform-neutral extensible mechanism for serializing structured data*”. This is based around so-called “*building blocks*” which are standardised fields that can be used to structure information. We adapted it to aggregate phenotype meta-analytic data. Specifically, the following changes were added to this framework: we added “*Publication*” as a new top-level field and took the PMID structured fields as building blocks for this entry. This process allowed us to fully automate field annotation by providing the .csv download data from Pubmed searches. In addition to this, we added several new “*building-blocks*” to the existing top-level field cohort. These changes/adaptations are summarised in table 1. Because published cohort studies may include several cohorts, we added a suffix the main top-level cohort field _[number], but where single case reports were extractable defaulted native to the phenopacket standard. We identified the distinction by labelling the outputs either “*phenopacket*” or “*metaphenome-annot”*. Regarding the misdiagnosis field, we identified two different *types* of misdiagnoses that may be described in a cohort study, depending on whether the analysis is looking forwards or backwards in time. The former involves a published cohort being labelled by their diagnosis in life but the post-mortem shows otherwise, which we label as a *prospective misdiagnosis*. In the latter case, a cohort may be identified at post-mortem with the same “gold standard” diagnosis, but review of the historical records reveals a different diagnosis in life, which we label a *retrospective misdiagnosis*. The distinction allows us to subsequently combine both types of data. We also added a pathology block, to capture reported histopathological data. This embedded Braak AD stage, ^2^ Thala Beta stage, ^3^ Plaque Score (qualitative), MSA pathological subtype, Lewy Body Disorder subtype and Likelihood of DLB into the main toolbox but could also manually define other schemes. Finally, we added a dual diagnosis field where mixed proteinopathies were clearly identified and labelled (however in the future this may be subsumed by the pathology field). All cohort data only included the total number with the post-mortem confirmed diagnoses (i.e., misdiagnosis data was excluded/subtracted from these figures at the time of annotation and entered separately in the misdiagnosis fields).

Because our objective was to produce machine readable files that also align with the Brain Imaging Dataset Standard ^4^ (BIDS, https://bids-specification.readthedocs.io/en/stable/), we chose .json as the default output. However, it is possible to convert these to protobuf where required. The resulting output filename was structured as follows:

- Metaphenome file: [PMID]_[FIRST-AUTHOR]_[PUB-DATE]_metaphenome-annot.json
- Phenopacket file: [PMID]_[FIRST-AUTHOR]_[PUB-DATE]_phenopacket-sub-[no].json

Where PMID is the pubmed ID, and publication date provided in the form yyyymmdd. Finally, for all summary statistics, if the median and confidence intervals were provided, they were also converted to mean and standard deviation using the method described by Wan et al 2014 to facilitate second-level data aggregation. ^5^

To note, since the analysis for this work was concluded there was a major update to the phenopacket schema which added their own “*measurement*” fields (absent in version 1.0 when changes in table 1 were specified). Whilst we have attempted to align as closely as possible with the original phenopacket schema, the metaphenomic implementation has been designed with a slightly different purpose in mind (i.e., optimised for cohort data aggregation rather than single case descriptions) and retrofitting the updated measurement field would require a substantial update to the toolbox and underlying code. Therefore, for the purposes of this work we have retained our original framework, and plan to harmonise these differences in future updates.

**Supplementary Table 1:**
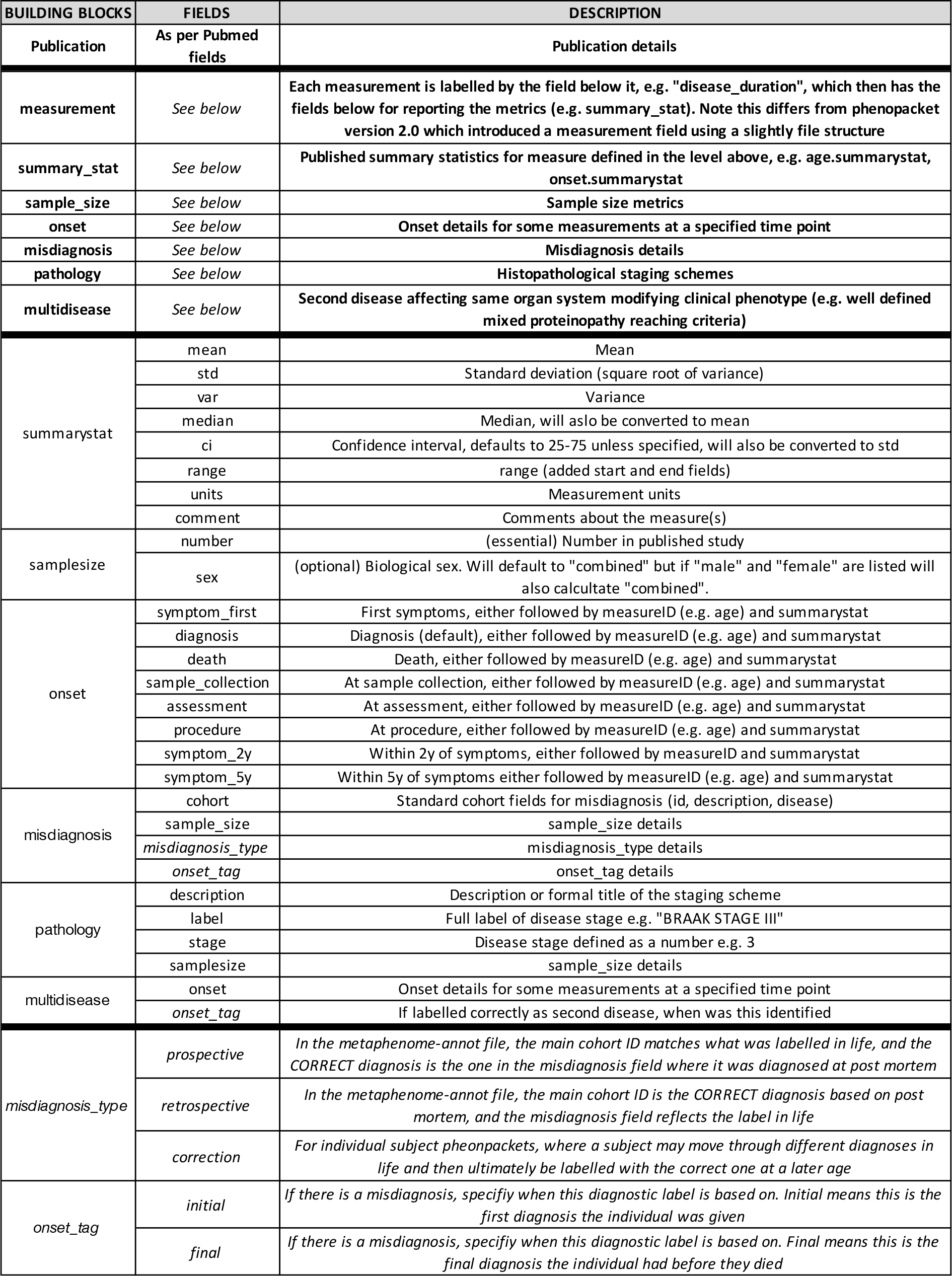
Summary of changes to the phenopacket schema.

### S1.2 Calculating metrics of diagnostic accuracy

In this work, we were interested in pooling information across different Parkinsonian disorders and temporally directed analyses (post-mortem compared to life-time diagnosis and visa versa) to try and provide more granular insights into overall diagnostic accuracy. To summarise these, we collapsed the data for each disease into a 2×2 confusion matrix, where the diagnosis in life was framed as the prediction, and pathological diagnosis as the ground truth. In this way, we could calculate the corresponding sensitivity, specificity and balanced accuracy for each clinical diagnosis, conditional on all of the main Parkinsonian disorders.

**Supplementary Figure 1:**
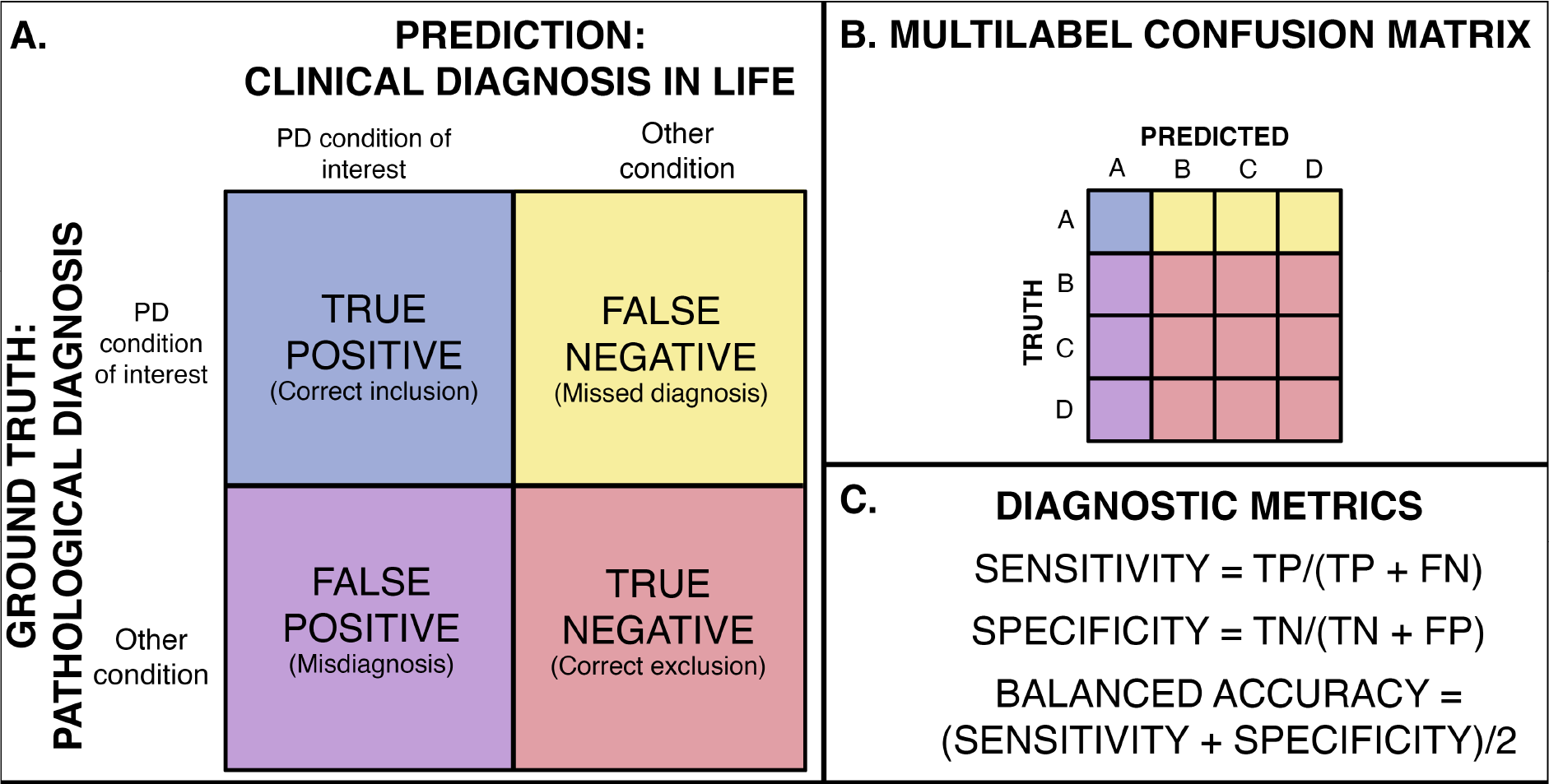
**A**. Diagnosis definitions mapped to 2×2 confusion matrix; **B**. Collapsing multiple categories into 2×2 confusion matrix to calculate summary metrics, in this example the disease of interest is “A” ; **C**. Calculation of diagnostic accuracy metrics.

### S1.3 Calculating probability of disease from phenotypic features

We adopted a naïve Bayesian classifier approach similar to the probability of prodromal Parkinson’s disease approach. ^6^ The advantage is that it allows diagnostic information to be sequentially added and used to update pretest probability of disease (P) given new information. Furthermore, providing our results in this format also means they can easily be used by other similar classifiers allowing *in vivo* models to incorporate diagnostic uncertainty and leverage post-mortem defined likelihood ratios (LR). Finally, by providing the original underlying data in a structured, reusable, machine-readable format, these probabilities can be rapidly and iteratively updated as new results emerge. This approach has been described elsewhere ^6^ but has been summarized below for clarity:

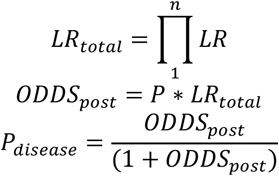

Where:

➢ P = Pre-test probability
➢ LR = Likelihood ratios for features of interest
➢ n = Total number of observed features with LRs
➢ LR_total_ = Pooled likelihood ratio
➢ ODDS_post_ = Post-test odds
➢ P_disease_ = Probability of disease

Because the objective was to provide a means to quantify diagnostic precision in an individual presenting with Parkinsonism, we did not incorporate the population prevalence into our model, but simply calculated our pre-test odds (P) based on age at presentation directly from the post-mortem data.

There was marked variability in the literature as to when an illness’ various features (phenotypic traits) were described, what was described or omitted, how they were categorized, and the ontologies used. For simplicity, in this analysis a phenotype was included if it occurred at any point during the illness, and the sample population was calculated for each phenotype by only counting studies where that feature was described. Overall, 88% of studies provided some data that could be extracted, providing 4076 features. After review, these could be collapsed to 246 unique HPO terms. This exercise resulted in a number of new terms and suggested modifications to the existing HPO framework improve coverage for these movement disorders. Supplementary figure 2 provides an example of this framework for the HPO term Parkinsonism:

**Supplementary Figure 2:**
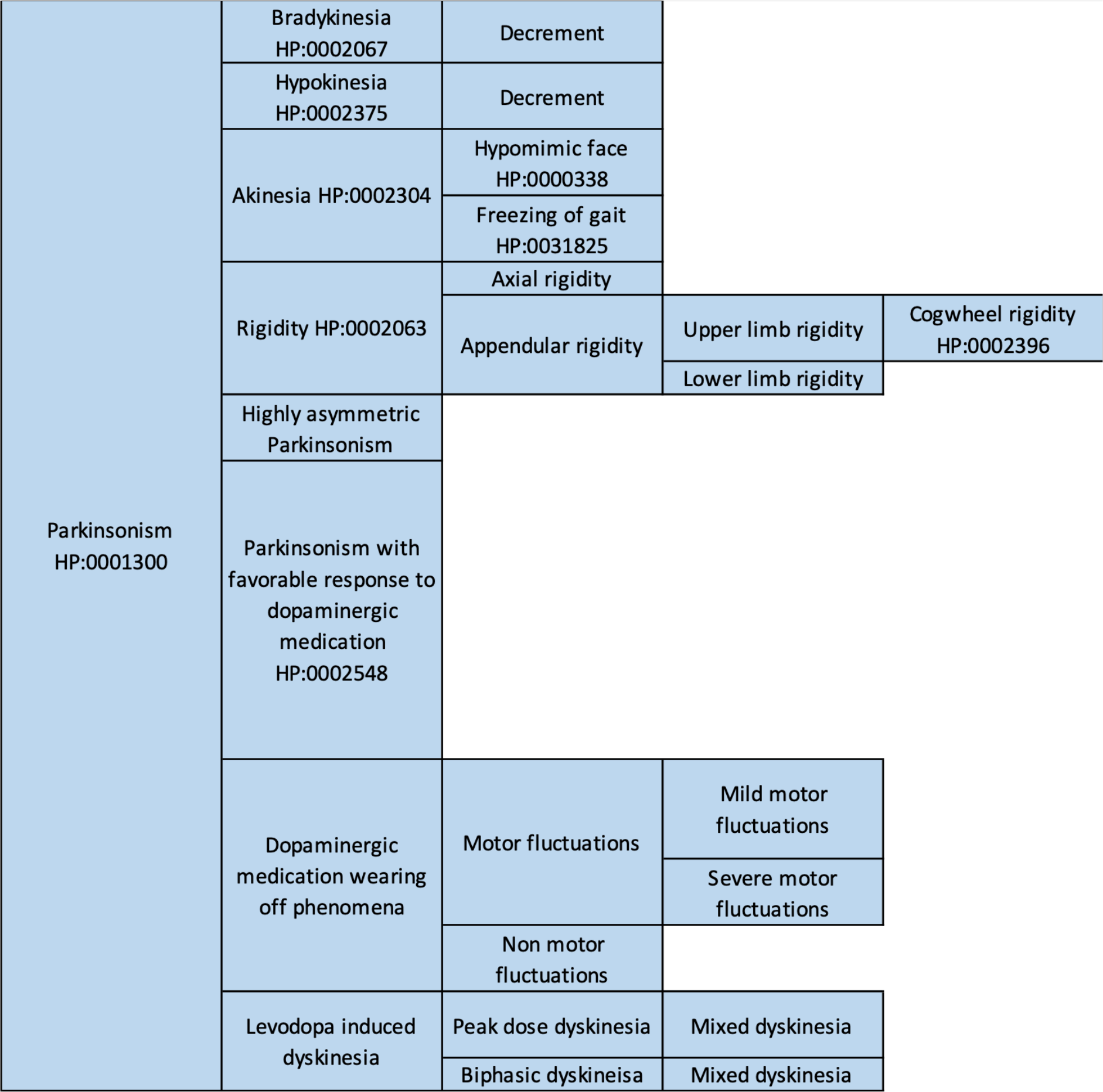
HPO Inheritance principle optimised for Parkinson’s disease. (part of Abnormality of movement HP:0100022 shown). Note all terms with missing HPO IDs have been defined and submitted to HPO to provide better coverage Parkinsonian for movement disorders.

Each unique HPO term was then mapped to the broader HPO hierarchical framework, which organises terms as a directed acyclic graph where each sub-term (child) represents a more specific or limited instance of its parent term(s) and can be connected by a *“is-a”* (>) relationship, for example:

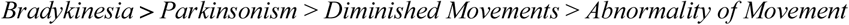

#### Bradykinesia > Parkinsonism > Diminished Movements > Abnormality of Movement

This means that incomplete coverage of low-frequency phenotypes still contribute to provide better coverage further up the HPO hierarchy. Because this approach combines both single case studies and group cohort data, propagating information up the HPO tree represents a challenge because, in contrast to single subject observations, a negative observation in one of the child terms does not necessarily mean the parent term was absent – Taking only the positive observations risked up-weighting rare phenotypes, whereas summing both the present and absent phenotypes present in all child terms risked down-weighting more common higher-level parent phenotypes. Because the objective was to create likelihood ratios of one disease relative to another, we were able to test both approaches by calculating the LR for every HPO term in the hierarchy, across all the diseases, and reviewing the top-ranked terms (Figure 7). We found summing both present and absent observations provided the best solution, provided observations were present for eight or more individuals for the main group, and applied this to the entire HPO tree to calculate positive and negative likelihood ratios for every phenotypic feature between the five main conditions (PD, MSA, PSP, DLB, CBD). We removed any LRs where the denominator was zero in this work (i.e., LR = infinity) – Whilst these may reflect pathognomonic clinical signs, given the stark variability in what clinical phenotypes were reported in the clinic-pathological literature, we could confidently rank these as such in this work, as some of the results were clearly artefactual due to under-reporting and incomplete coverage. This limitation could be offset in the future by incorporating more detailed observations from other cohorts (e.g., *in vivo* observational studies).

### S1.4 Data visualisation

All data visualisation code was created by CL and implemented in MATLAB - Gaussian probability density, cumulative distribution functions and heatplots were generated using inbuilt MATLAB functions. The RainCloud plots were adapted from Allen. ^7^. The riverplots used to summarise change in diagnosis were adapted from the “Sankey Diagram” code available via MATLAB central. ^8^ The colorbrewer palattes were used. ^9^ All data-visualisation code has been made available with the main analysis code at: (url)

## S2. Supplementary Results

### S2.1 List of annotated clinicopathological studies

1, Sobue G,1992: Somatic motor efferents in multiple system atrophy with autonomic failure: a clinico-pathological study

2, Hughes AJ,1992: Accuracy of clinical diagnosis of idiopathic Parkinson’s disease: a clinico-pathological study of 100 cases

3, Jankovic J,1993: What is it? Case 1, 1993: parkinsonism, dysautonomia, and ophthalmoparesis

4, Churchyard A,1993: Dopa resistance in multiple-system atrophy: loss of postsynaptic D2 receptors

5,,1993: Case records of the Massachusetts General Hospital. Weekly clinicopathological exercises. Case 46-1993. A 75-year-old man with right-sided rigidity, dysarthria, and abnormal gait

6, Wenning GK,1994: Clinical features and natural history of multiple system atrophy. An analysis of 100 cases

7, Harris CP,1994: Depression followed by dementia and disordered movement. Clinicopathologic correlation

8, de Vos RA,1995: ‘Lewy body disease’: clinico-pathological correlations in 18 consecutive cases of Parkinson’s disease with and without dementia

9, Wenning GK,1995: Clinicopathological study of 35 cases of multiple system atrophy

10, Collins SJ,1995: Progressive supranuclear palsy: neuropathologically based diagnostic clinical criteria

11, Litvan I,1996: Validity and reliability of the preliminary NINDS neuropathologic criteria for progressive supranuclear palsy and related disorders

12, Litvan I,1996: Natural history of progressive supranuclear palsy (Steele-Richardson-Olszewski syndrome) and clinical predictors of survival: a clinicopathological study

13, Verny M,1996: Progressive supranuclear palsy: a clinicopathological study of 21 cases 14, Bergeron C,1996: Unusual clinical presentations of cortical-basal ganglionic degeneration

15, Litvan I,1997: Which clinical features differentiate progressive supranuclear palsy (Steele-Richardson-Olszewski syndrome) from related disorders? A clinicopathological study

16, Schneider JA,1997: Corticobasal degeneration: neuropathologic and clinical heterogeneity

17,,1997: Case records of the Massachusetts General Hospital. Weekly clinicopathological exercises. Case 26-1997. A 64-year-old man with progressive dementia, seizures, and unstable gait

18, Tsuchiya K,1997: Distribution of cerebral cortical lesions in corticobasal degeneration: a clinicopathological study of five autopsy cases in Japan

19, Wenning GK,1998: Natural history and survival of 14 patients with corticobasal degeneration confirmed at postmortem examination

20, Boeve BF,1999: Pathologic heterogeneity in clinically diagnosed corticobasal degeneration

21, Wenning GK,1999: Time course of symptomatic orthostatic hypotension and urinary incontinence in patients with postmortem confirmed parkinsonian syndromes: a clinicopathological study

22, Wenning GK,1999: Progression of falls in postmortem-confirmed parkinsonian disorders

23, Grimes DA,1999: Dementia as the most common presentation of cortical-basal ganglionic degeneration 24, Litvan I,1999: Clinicopathologic case report. Dementia with Lewy bodies (DLB)

25, Hohl U,2000: Diagnostic accuracy of dementia with Lewy bodies

26, Armstrong RA,2000: A quantitative study of the pathological lesions in the neocortex and hippocampus of twelve patients with corticobasal degeneration

27, Tsuchiya K,2000: Constant involvement of the Betz cells and pyramidal tract in multiple system atrophy: a clinicopathological study of seven autopsy cases

28, Mimura M,2001: Corticobasal degeneration presenting with nonfluent primary progressive aphasia: a clinicopathological study

29, Müller J,2001: Progression of dysarthria and dysphagia in postmortem-confirmed parkinsonian disorders

30, Mann DM,2001: Anosmia in dementia is associated with Lewy bodies rather than Alzheimer’s pathology

31, Vitaliani R,2002: The pathology of the spinal cord in progressive supranuclear palsy

32, Hughes AJ,2002: The accuracy of diagnosis of parkinsonian syndromes in a specialist movement disorder service

33, Müller J,2002: Freezing of gait in postmortem-confirmed atypical parkinsonism

34, Josephs KA,2002: A clinicopathological study of vascular progressive supranuclear palsy: a multi-infarct disorder presenting as progressive supranuclear palsy

35, Harding AJ,2002: Clinical correlates of selective pathology in the amygdala of patients with Parkinson’s disease

36, Poewe W,2002: The differential diagnosis of Parkinson’s disease

37, Birdi S,2002: Progressive supranuclear palsy diagnosis and confounding features: report on 16 autopsied cases 38, Mochizuki A,2003: Progressive supranuclear palsy presenting with primary progressive aphasia--clinicopathological report of an autopsy case

39, Colosimo C,2003: Lewy body cortical involvement may not always predict dementia in Parkinson’s disease

40, Osaki Y,2004: Accuracy of clinical diagnosis of progressive supranuclear palsy

41, Lezcano E,2004: Parkinson’s disease-like presentation of multiple system atrophy with poor response to STN stimulation: a clinicopathological case report

42, Schlossmacher MG,2004: Case records of the Massachusetts General Hospital. Weekly clinicopathological exercises. Case 27-2004. A 79-year-old woman with disturbances in gait, cognition, and autonomic function

43, Ozawa T,2004: The spectrum of pathological involvement of the striatonigral and olivopontocerebellar systems in multiple system atrophy: clinicopathological correlations

44, Tsuchiya K,2005: Constant and severe involvement of Betz cells in corticobasal degeneration is not consistent with pyramidal signs: a clinicopathological study of ten autopsy cases

45, Josephs KA,2005: Extending the clinicopathological spectrum of neurofilament inclusion disease

46, Williams DR,2005: Characteristics of two distinct clinical phenotypes in pathologically proven progressive supranuclear palsy: Richardson’s syndrome and PSP-parkinsonism

47, Tsuboi Y,2005: Increased tau burden in the cortices of progressive supranuclear palsy presenting with corticobasal syndrome

48, Halliday GM,2005: A comparison of degeneration in motor thalamus and cortex between progressive supranuclear palsy and Parkinson’s disease

49, Papapetropoulos S,2005: Natural history of progressive supranuclear palsy: a clinicopathologic study from a population of brain donors

50, Josephs KA,2005: Atypical progressive supranuclear palsy underlying progressive apraxia of speech and nonfluent aphasia

51, Josephs KA,2006: Clinicopathological and imaging correlates of progressive aphasia and apraxia of speech 52, Kłodowska-Duda G,2006: Corticobasal degeneration -- clinico-pathological considerations

53, Murray R,2007: Cognitive and motor assessment in autopsy-proven corticobasal degeneration

54, Kempster PA,2007: Patterns of levodopa response in Parkinson’s disease: a clinico-pathological study

55, Compta Y,2007: Long lasting pure freezing of gait preceding progressive supranuclear palsy: a clinicopathological study

56, Facheris MF,2008: Pure akinesia as initial presentation of PSP: a clinicopathological study

57, Jellinger KA,2008: Different tau pathology pattern in two clinical phenotypes of progressive supranuclear palsy

58, Lladó A,2008: Clinicopathological and genetic correlates of frontotemporal lobar degeneration and corticobasal degeneration

59, O’Sullivan SS,2008: Clinical outcomes of progressive supranuclear palsy and multiple system atrophy

60, Kalaitzakis ME,2009: Dementia and visual hallucinations associated with limbic pathology in Parkinson’s disease

61, Brooks D,2009: Intralaminar nuclei of the thalamus in Lewy body diseases

62, Kanazawa M,2009: Cerebellar involvement in progressive supranuclear palsy: A clinicopathological study 63, Rajput AH,2009: Course in Parkinson disease subtypes: A 39-year clinicopathologic study

64, Selikhova M,2009: A clinico-pathological study of subtypes in Parkinson’s disease

65, Sabbagh MN,2009: Parkinson disease with dementia: comparing patients with and without Alzheimer pathology

66, Molano J,2010: Mild cognitive impairment associated with limbic and neocortical Lewy body disease: a clinicopathological study

67, Kempster PA,2010: Relationships between age and late progression of Parkinson’s disease: a clinico-pathological study

68, Ozawa T,2010: The phenotype spectrum of Japanese multiple system atrophy

69, Ling H,2010: Does corticobasal degeneration exist? A clinicopathological re-evaluation

70, Espay AJ,2011: Rapidly progressive atypical parkinsonism associated with frontotemporal lobar degeneration and motor neuron disease

71, Snowden JS,2011: The clinical diagnosis of early-onset dementias: diagnostic accuracy and clinicopathological relationships

72, Kouri N,2011: Neuropathological features of corticobasal degeneration presenting as corticobasal syndrome or Richardson syndrome

73, Iodice V,2012: Autopsy confirmed multiple system atrophy cases: Mayo experience and role of autonomic function tests

74, Magdalinou NK,2013: Normal pressure hydrocephalus or progressive supranuclear palsy? A clinicopathological case series

75, Iwasaki Y,2013: An autopsied case of progressive supranuclear palsy presenting with cerebellar ataxia and severe cerebellar involvement

76, Boeve BF,2013: Clinicopathologic correlations in 172 cases of rapid eye movement sleep behavior disorder with or without a coexisting neurologic disorder

77, Shim YS,2013: Clinicopathologic study of Alzheimer’s disease: Alzheimer mimics

78, Men šíková K,2013: Progressive supranuclear palsy phenotype mimicking synucleinopathies

79, Fujioka S,2013: Similarities between familial and sporadic autopsy-proven progressive supranuclear palsy 80, Joutsa J,2014: Diagnostic accuracy of parkinsonism syndromes by general neurologists

81, Figueroa JJ,2014: Multiple system atrophy: prognostic indicators of survival

82, Adler CH,2014: Low clinical diagnostic accuracy of early vs advanced Parkinson disease: clinicopathologic study

83, Jacobson SA,2014: Plaques and tangles as well as Lewy-type alpha synucleinopathy are associated with formed visual hallucinations

84, Ikeda C,2014: Corticobasal degeneration initially developing motor versus non-motor symptoms: a comparative clinicopathological study

85, Respondek G,2014: The phenotypic spectrum of progressive supranuclear palsy: a retrospective multicenter study of 100 definite cases

86, Zhu MW,2015: Typical or atypical progressive supranuclear palsy: a comparative clinicopathologic study of three Chinese cases

87, Cykowski MD,2015: Expanding the spectrum of neuronal pathology in multiple system atrophy 88, Koga S,2015: When DLB, PD, and PSP masquerade as MSA: an autopsy study of 134 patients

89, Virmani T,2015: Clinicopathological characteristics of freezing of gait in autopsy-confirmed Parkinson’s disease

90, Iacono D,2015: Parkinson disease and incidental Lewy body disease: Just a question of time?

91, Xie T,2015: Comparison of clinical features in pathologically confirmed PSP and MSA patients followed at a tertiary center

92, Beach TG,2016: Prevalence of Submandibular Gland Synucleinopathy in Parkinson’s Disease, Dementia with Lewy Bodies and other Lewy Body Disorders

93, Koga S,2016: Cerebellar ataxia in progressive supranuclear palsy: An autopsy study of PSP-C

94, Kurz C,2016: An autopsy-confirmed case of progressive supranuclear palsy with predominant postural instability

95, Adamowicz DH,2017: Hippocampal α-Synuclein in Dementia with Lewy Bodies Contributes to Memory Impairment and Is Consistent with Spread of Pathology

96, Rajput AH,2017: Octogenarian parkinsonism - Clinicopathological observations

97, Walker L,2017: Quantitative neuropathology: an update on automated methodologies and implications for large scale cohorts

98, Suemoto CK,2017: Neuropathological diagnoses and clinical correlates in older adults in Brazil: A cross-sectional study

99, Turcano P,2017: Clinicopathologic discrepancies in a population-based incidence study of parkinsonism in olmsted county: 1991-2010

100, Jung Y,2018: Clinicopathological and (123)I-FP-CIT SPECT correlations in patients with dementia

101, Roudil J,2018: Influence of Lewy Pathology on Alzheimer’s Disease Phenotype: A Retrospective Clinico-Pathological Study

102, De Pablo-Fernández E,2019: Prognosis and Neuropathologic Correlation of Clinical Subtypes of Parkinson Disease

103, Stejskalova Z,2019: Pyramidal system involvement in progressive supranuclear palsy - a clinicopathological correlation

104, Miki Y,2019: Improving diagnostic accuracy of multiple system atrophy: a clinicopathological study

105, Jabbari E,2019: The genetic and clinico-pathological profile of early-onset progressive supranuclear palsy

106, Vergouw LJM,2020: Dementia With Lewy Bodies: A Clinicopathologic Series of False-positive Cases

107, Jabbari E,2020: Diagnosis Across the Spectrum of Progressive Supranuclear Palsy and Corticobasal Syndrome

108, Knox MG,2020: Neuropathological Findings in Parkinson’s Disease With Mild Cognitive Impairment

109, Koga S,2020: Clinicopathologic and genetic features of multiple system atrophy with Lewy body disease

110, Smirnov DS,2020: Cognitive decline profiles differ in Parkinson disease dementia and dementia with Lewy bodies

111, Koga S,2020: Cerebrovascular pathology and misdiagnosis of multiple system atrophy: An autopsy study

112, Homma T,2020: Cerebral white matter tau-positive granular glial pathology as a characteristic pathological feature in long survivors of multiple system atrophy

113, Boes S,2020: Dementia with Lewy bodies presenting as Logopenic variant primary progressive Aphasia

114, Longardner K,2020: Orthostatic hypotension preceding dementia with Lewy bodies by over 15 years: a clinicopathologic case report

115, Hansen D,2021: Novel clinicopathological characteristics differentiate dementia with Lewy bodies from Parkinson’s disease dementia

116, Ishida C,2021: Effectiveness of Levodopa in Patients with Multiple System Atrophy and Associated Clinicopathological Features

117, Zhang S,2021: Case Report of a pathologically confirmed vascular parkinsonism with early cognitive impairment and Behavioral disturbance

118, Homma T,2021: Digital mapping of Lewy bodies and neurites in alpha-synuclein stained large cerebral hemispheric sections from three patients with dementia with Lewy bodies showing psychotic manifestations: A pilot study

119, Crosiers D,2021: Cerebellar ataxia in progressive supranuclear palsy: a clinico-pathological case report

120, Chatterjee A,2021: Clinico-pathological comparison of patients with autopsy-confirmed Alzheimer’s disease, dementia with Lewy bodies, and mixed pathology

121, Donlon E,2021: Braak’s Unfinished Hypothesis: A Clinicopathological Case Report of α-Synuclein Peripheral Neuropathy Preceding Parkinsonism by 20 Years

122, Kawakatsu S,2021: Clinicopathological heterogeneity of Alzheimer’s disease with pure Alzheimer’s disease pathology: Cases associated with dementia with Lewy bodies, very early-onset dementia, and primary progressive aphasia

123, Natera-Villalba E,2021: Eye-of-the-Tiger Sign with an Unexpected Pathological Diagnosis 124, Lin CR,2022: Clinicopathological correlates of pyramidal signs in multiple system atrophy

125, Horimoto Y,2022: A descriptive study of Parkinson disease and atypical parkinsonisms in the Annuals of the Pathological Autopsy Cases in Japan

